# Surgical versus Non-Surgical Treatment for Esophageal Squamous Cell Carcinoma in Patients Older than 70 years: A Propensity Score Matching Analysis

**DOI:** 10.1101/2023.08.01.23293469

**Authors:** Kexun Li, Changding Li, Xin Nie, Wemwu He, Kunyi Du, Kun Liu, Chenghao Wang, Jialong Li, Kunhan Ni, Yongtao Han, Lin Peng, Qifeng Wang, Xuefeng Leng

## Abstract

**Background:** With the rising life expectancy and an ageing population, it has become increasingly important to investigate treatments suitable for older adult patients with esophageal cancer. This study investigated whether older adult patients who underwent esophagectomy had better clinical outcomes than those who were non-surgically treated.

**Materials and methods:** We retrospectively analyzed patients with esophageal squamous cell carcinoma (ESCC) who were 70 years or older and underwent esophagectomy, radiotherapy (RT), and/or chemotherapy (CT) from January 2018 to December 2019. Patients were divided into two groups: the surgery group (S group) and non-surgery group (NS group). Then compared the clinical outcomes of the two groups.

**Results:** After a median follow-up duration of 36.6 months, the S group showed better OS. The 3-year OS was 59% in the S group and 27% in the NS group (HR, 0.397; 95% CI, 0.278–0.549; P<0.0001). In the S group, the median progression free survival was 38.3 months (95% CI, 30.6–46.1) compared to 12.3 months in the NS group (HR, 0.511; 95% CI, 0.376–0.695; P<0.0001). In addition, the number of adverse events in the NS group was higher than that in the S group (P<0.001)

**Conclusion:** Overall, patients with ESCC ≥70 years who underwent esophagectomy had significantly better clinical outcomes than those who underwent non-surgical treatment with RT and/or CT.

- **What is already known on this topic** – There was a lack of comprehensive research specifically focusing on older adult patients with ESCC and comparing the outcomes of surgical and non-surgical treatments. Given the rising life expectancy and aging population, it became increasingly important to investigate appropriate treatment options for older adult patients with esophageal cancer. This study aimed to fill the gap in knowledge by retrospectively analyzing patients with ESCC who were 70 years or older and underwent different treatment modalities such as esophagectomy, radiotherapy (RT), and/or chemotherapy (CT).By conducting this study, the researchers aimed to provide valuable insights into the optimal treatment approach for older adult patients with ESCC. Understanding the comparative effectiveness and safety of surgical versus non-surgical treatments would help clinicians make informed decisions when managing and treating this specific patient population.
- **What this study adds** – In addition to the research discussion on OS and DFS, this study also increased the comparison of Adverse events of different treatments in elderly ESCC patients.
- **How this study might affect research, practice or policy** – In the treatment practice of elderly patients with ESCC, it is suggested that surgical treatment is possible.

## 1. INTRODUCTION

According to the Global Cancer Statistics 2020: GLOBOCAN Estimates of Incidence and Mortality Worldwide for 36 Cancers in 185 Countries, the incidence of esophageal cancer ranked seventh among the 36 human cancers and was the sixth primary cause of cancer-related death in the world, with the highest incidence rates for both men and women seen in Eastern Asia, particularly China [1]. Esophageal squamous cell carcinoma (ESCC) is a significant disease impacting the Chinese population’s health and overall survival (OS). In recent years, given the increase in the number of studies on ESCC, comprehensive treatment modalities based on esophagectomy have gradually improved [2–5].

Based on the analysis of lifetime death probability from major causes of death among Chinese residents, heart diseases, cerebrovascular diseases, malignant tumors, respiratory diseases, and injuries from poisoning are the five major causes of death in the Chinese population [6]. Among the aforementioned conditions, cancer is the most high-risk cause of death in people aged 40–45 years, while death in older adult patients is mostly related to cardiovascular and cerebrovascular diseases [1]. The median age of diagnosis for esophageal cancer is 68, and more than 40 percent of patients are over 70 years [7,8]. With the rising life expectancy and an ageing population, it has become increasingly important to investigate treatments suitable for older patients with esophageal cancer. In China, the number of patients with ESCC aged 70 years or older who visited the Sichuan Cancer Hospital has increased yearly, from only 18 cases in 2010 to 139 cases in 2016.

Esophagectomy is performed through a right transthoracic procedure with two or three-field lymph node dissections [9–12]. This procedure, known as McKeown or Ivor-Lewis esophagectomy, has been the main surgical approach used in patients with ESCC. However, elderly patients may be more susceptible to postoperative complications such as infections, delirium, and cognitive dysfunction, which can lead to longer hospital stays, increased healthcare costs, and decreased quality of life [13–15]. In addition, studies have shown that elderly patients with ESCC have higher short-term mortality and lower long-term survival after esophagectomy [16]. Currently, there remains a lack of high-quality clinical evidence that compares surgical versus non-surgical treatment modalities for patients with ESCC in China, especially for the older adult population aged 70 or older. In this study, we aimed to assess whether older adult patients aged 70 or above are suitable for comprehensive treatment, that includes a surgical intervention, through prospective verification.

## 2. MATERIALS AND METHODS

### 2.1. Study design

We collected data from a retrospective database (Sichuan Cancer Hospital & Institute Esophageal Cancer Case Management Database (SCH-ECCM Database)) on 284 patients with ESCC from a cancer hospital between January 2018 to December 2019. The patients were divided into two groups: the surgery group (S group) and the non-surgery group (NS group). We then measured the patients’ clinical outcomes. Furthermore, patients’ records were reviewed for clinicopathologic findings and outcomes. Esophagectomy was performed through a right transthoracic procedure with a two-field or three-field lymphadenectomy. The surgical approach used depended on the characteristics of the patient and the surgeon’s preferences. Disease staging was adjusted to the American Joint Committee on Cancer/Union for International Cancer Control 8th edition tumor–node–metastasis (TNM) system. There were three inclusion criteria: (1) patients with pathologically confirmed diagnosis of ESCC, (2) patients who underwent esophagectomy, RT, and/or CT, and (3) patient’s age was ≥70 years. There were two exclusion criteria: (1) the presence of other malignant tumors and (2) missing required data. Patients were then assigned to the S or NS group (Figure 1). From 2018 to 2019, paclitaxel in combination with platinum was the primary regimen, with a small number of patients receiving Tegafur or docetaxel in combination with platinum. If a patient was placed into the NS group but salvage esophagectomy was later performed, that patient’s outcomes were excluded. The patients’ records were prospectively collected along with data regarding clinicopathologic findings and outcomes. Patients were followed up once every 3 months for the first 2 years, and once every 6 months after that. OS was calculated from the month and year of initial diagnosis till death or last follow-up, while progression-free survival (PFS) was recorded from the date of treatment/surgery until detection of recurrence, progression, or metastasis. The last follow-up was in April 2022. All procedures performed in this study were in accordance with the Declaration of Helsinki (as revised in 2013). This study was approved by the Ethics Committee of the Sichuan Cancer Hospital (No. NCC2014ZC-01) and Sichuan Science and Technology Project (No. 2021YJ0118) and this retrospective study was reviewed by the Ethics Committee (EC) for Medical Research and New Medical Technology of Sichuan Cancer Hospital (SCCHEC-02-2022-050). The data are anonymous, and the requirement for informed consent was therefore waived. The work has been reported in line with the STROBE criteria [17].

**Figure 1.** CONSORT diagram showing patient selection. ESCC, esophageal squamous cell carcinoma.

### 2.2. Criteria and characteristics of the adverse events

Based on relevant reports and international standards from the World Health Organization (WHO), body mass index (BMI) was divided into three levels [18,19]. According to the recommendation of the International Consensus on Standardization of Data Collection for Complications Associated with Esophagectomy [20], the Clavien–Dindo classification was used as a reference guide to standardize the classification of surgical complications [21–23]. Toxicities after CT, RT, and chemoradiotherapy (CRT) were evaluated using the common terminology criteria, Common Terminology Criteria for Adverse Events (CTCAE), for adverse events [24,25].

### 2.3. Statistical analysis

OS and PFS curves were calculated using the Kaplan–Meier method and compared using the unstratified log-rank test. A Cox regression model with stepwise selection was used for multivariate analyses, and the results are reported as hazard ratios (HRs) with 95% confidence intervals (CIs). P-values of <0.05 were considered statistically significant. All analyses were performed using SPSS version 23.0 (SPSS Inc. Chicago, IL, USA). Furthermore, unbalanced covariates were adjusted by performing propensity score matching (PSM) to create two comparable groups of patients: Group S and Group NS. A logit model was used to estimate patient propensity scores that included age, sex, Karnofsky Performance Status (KPS) scores, cT stage, cN stage, TNM stage, tumor location, BMI, and comorbidity incorporation for the cohort of PSM. Nearest neighbor matching (1:1) was performed without replacement based on a prespecified caliper width (0.02) to match patients in the NS and S groups. Patients with TNM stage I and patients with a missing tumor location during PSM were excluded.

## 3. RESULTS

### 3.1. Patient characteristics

We enrolled 284 patients in this study, 148 of whom (52.1%) underwent surgery while 136 (47.9%) underwent non-surgical treatment. Approximately 85.6% (243/284) of the patients were between 70–79 years of age, and 14.4% (41/284) were 80 years old or above; 79.9% (227/284) were men, while 20.1% (57/284) were women. Patients with clinical stage III accounted for 49.3%. The proportion of patients with clinical stage N0 was 13.7% (39/284) versus 86.3% (245/284) of those with clinical N+ stage. Of the patients, 85.4% (243/284) had a BMI below 18.5, and 40.5% had other comorbidities: 27.5% had high blood pressure (HBP), 8.5% had diabetes mellitus (DM), 3.9% had coronary heart disease (CHD), and 9.2% had chronic obstructive pulmonary disease (COPD) (Table 1). The incidence of non-R0 resection was 3.4% (5/148).

**Table 1.**
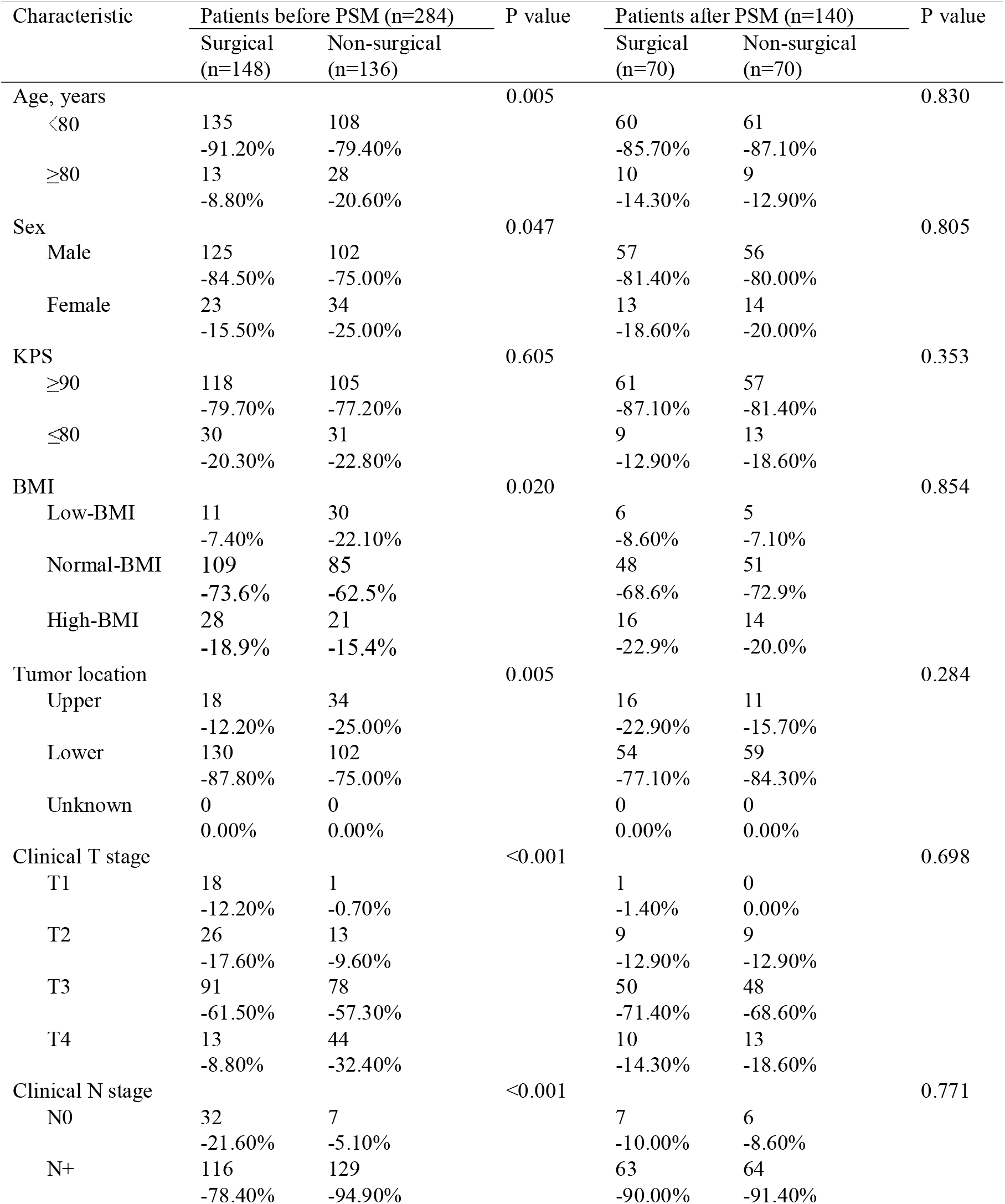
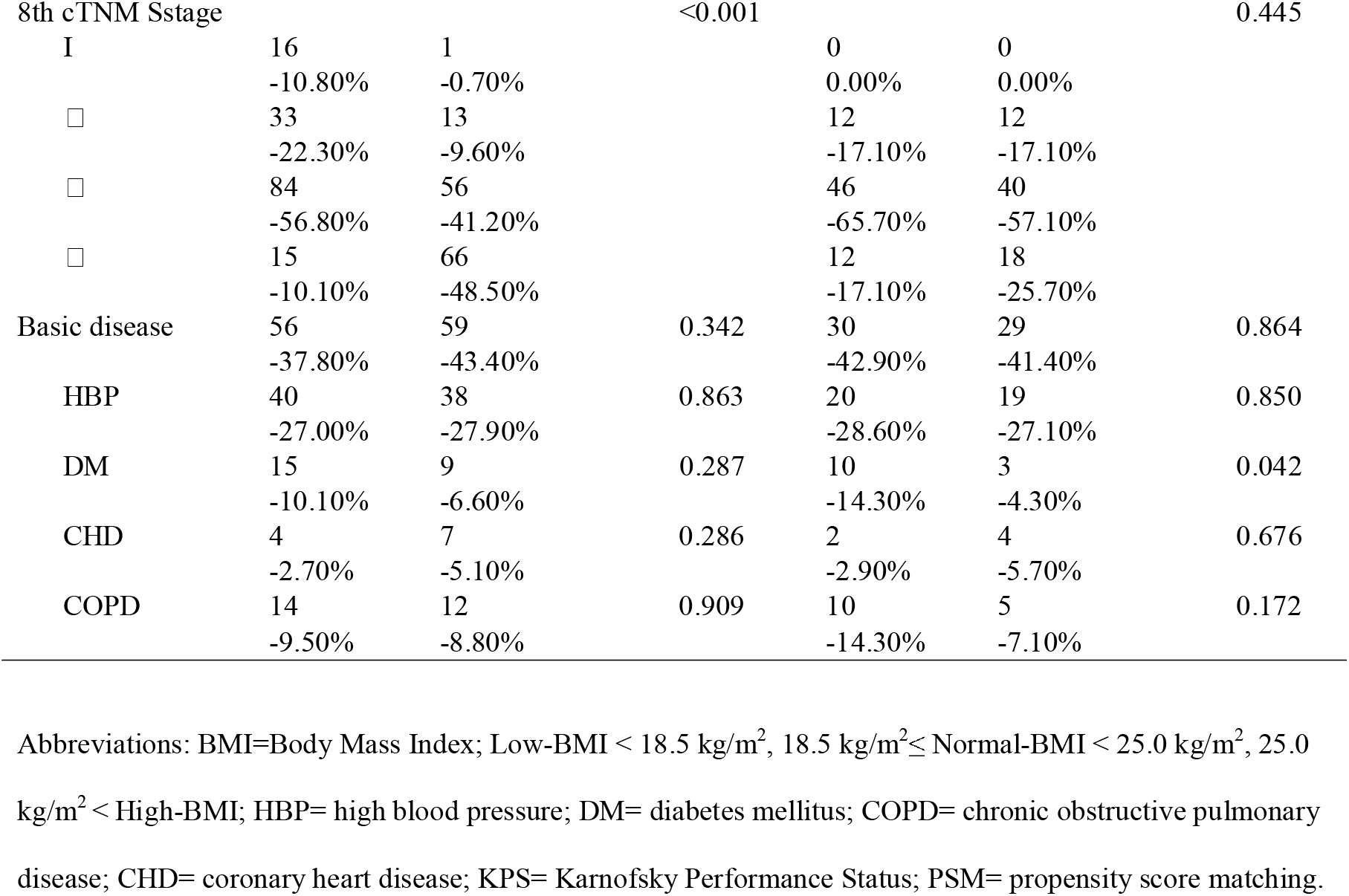
Demographic characteristics of the patients.

### 3.2. Survival outcomes

After 36.6 months of median follow-up time, the median OS is 24.4 months in 284 patients (95% CI, 17.3–31.6). In the S group, OS did not reach the median overall survival time, with the average OS being 35.1 months (95% CI, 32.0–38.2), while the median OS of the NS group was only 15.4 months (95% CI, 9.2–21.6). The OS at 1, 2, and 3 years was 80%, 66%, and 59% in the S group, respectively. In the NS group, the OS rates at 1, 2, and 3 years were 54%, 34%, and 27%, respectively (HR, 0.397; 95% CI, 0.278–0.549; P<0.0001; Figure 2A). In the S group, the median PFS was 38.3 months (95% CI, 30.6–46.1) compared to 12.3 months in the NS group (HR, 0.511; 95% CI, 0.376–0.695; P<0.0001; Figure 2B). After excluding patients who died within 90 days of treatment, the OS of the S group was significantly higher than that of the NS group (HR, 0.681; 95% CI, 0.550–0.843; P=0.0001; Supplementary Figure 4A).

**Figure 2.** Overall survival curves before and after PSM. (A) The overall survival curves for the S group and NS group; (B) The progression-free survival curves for the S group and NS group; (C) The overall survival curve for the S group and NS group after PSM; (D) The progression-free survival curve for the S group and NS group after PSM. PSM, propensity score matching.

### 3.3. Subgroup analysis according to patient characteristics

The univariate analysis indicated that substantial factors influenced the 3-year OS after treatment, such as age (P=0.031), KPS (P=0.005), treatment method (P<0.001). Further analysis using multivariate methods indicated that the significant factors that affected the 3-year OS after treatment were the KPS (P=0.004), cTNM staging (P<0.001), and treatment method (P=0.005) (Table 2).

**Table 2.**
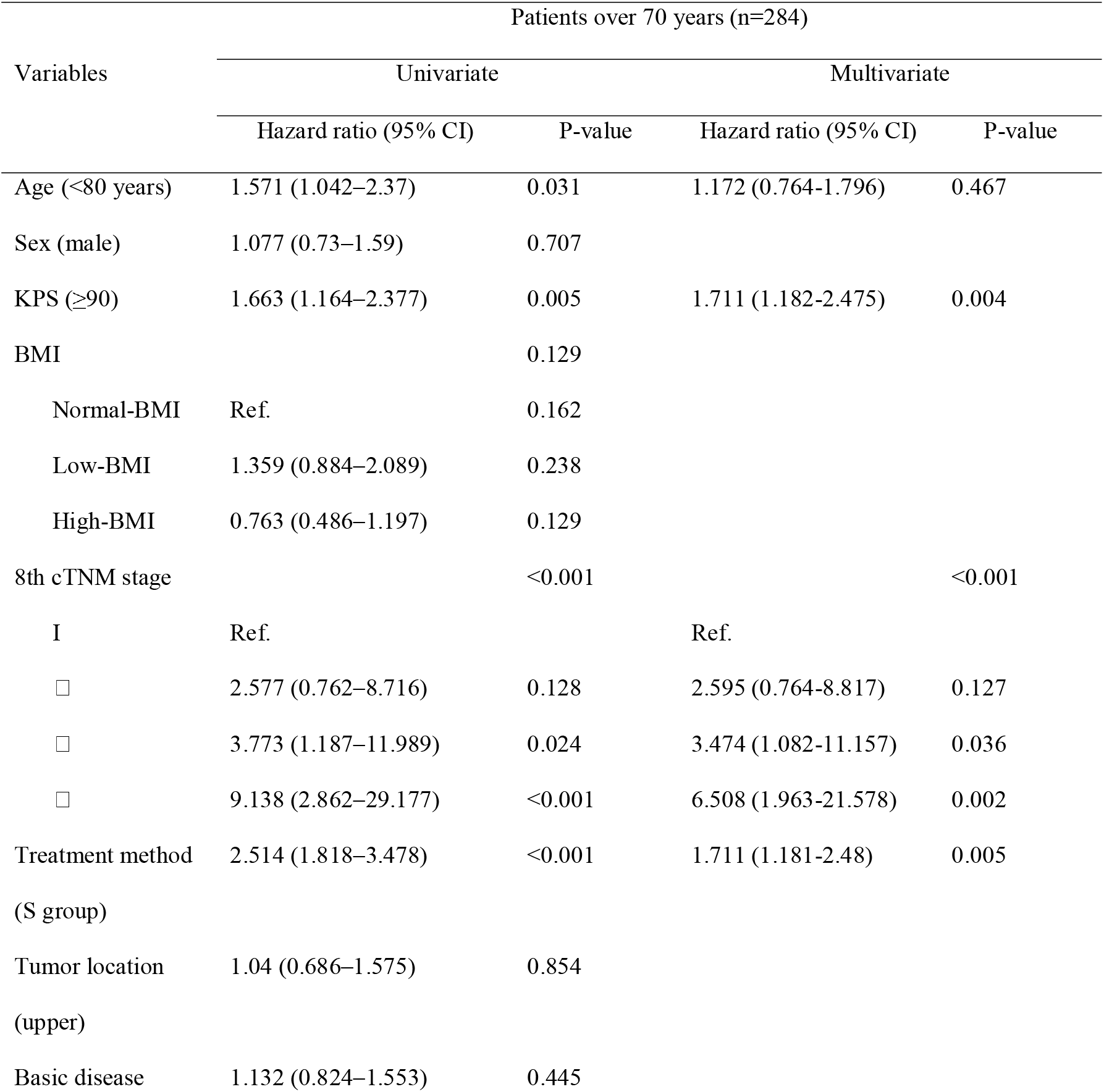
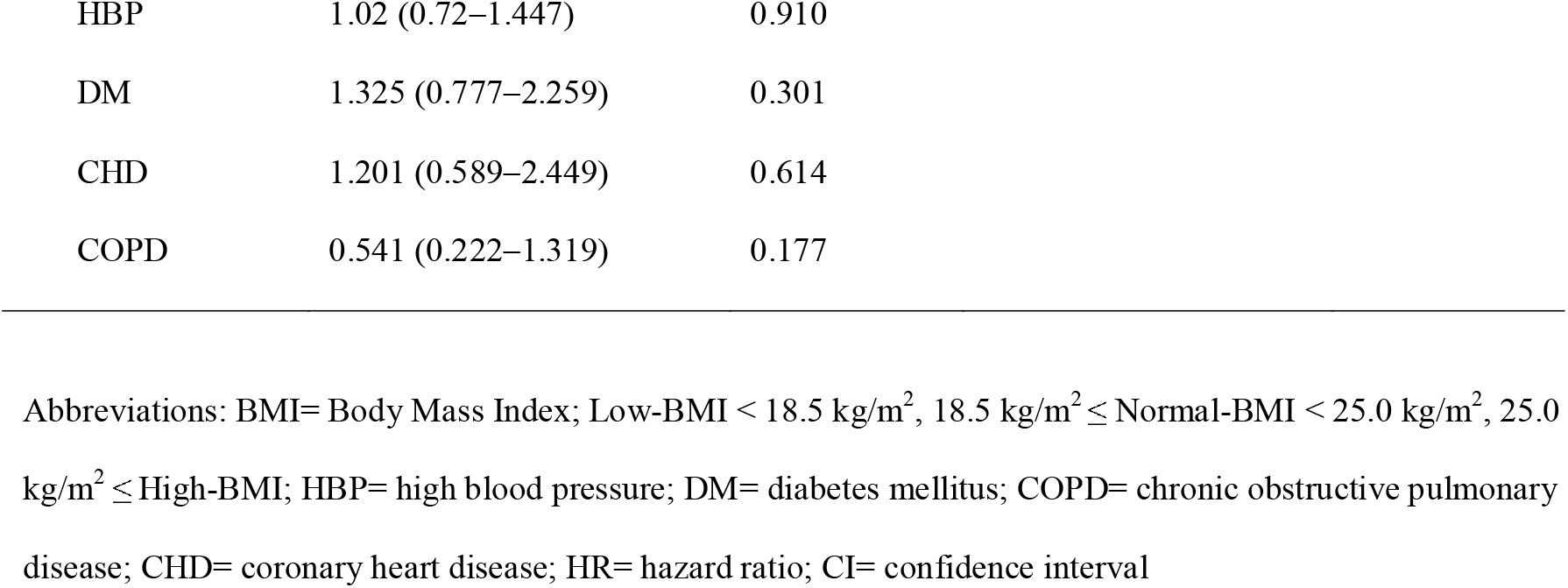
Univariate and multivariate Cox regression analysis factors affecting survival among patients.

| Variables | Patients over 70 years (n=284) |  |  |  |
| --- | --- | --- | --- | --- |
|  | Univariate |  | Multivariate |  |
|  | Hazard ratio (95% CI) | P-value | Hazard ratio (95% CI) | P-value |
| Age (<80 years) | 1.571 (1.042–2.37) | 0.031 | 1.172 (0.764-1.796) | 0.467 |
| Sex (male) | 1.077 (0.73–1.59) | 0.707 |  |  |
| KPS ($\geq 90$) | 1.663 (1.164–2.377) | 0.005 | 1.711 (1.182-2.475) | 0.004 |
| BMI |  | 0.129 |  |  |
| Normal-BMI | Ref. | 0.162 |  |  |
| Low-BMI | 1.359 (0.884–2.089) | 0.238 |  |  |
| High-BMI | 0.763 (0.486–1.197) | 0.129 |  |  |
| 8th cTNM stage |  | <0.001 |  | <0.001 |
| I | Ref. |  | Ref. |  |
| □ | 2.577 (0.762–8.716) | 0.128 | 2.595 (0.764-8.817) | 0.127 |
| □ | 3.773 (1.187–11.989) | 0.024 | 3.474 (1.082-11.157) | 0.036 |
| □ | 9.138 (2.862–29.177) | <0.001 | 6.508 (1.963-21.578) | 0.002 |
| Treatment method | 2.514 (1.818–3.478) | <0.001 | 1.711 (1.181-2.48) | 0.005 |
| (S group) |  |  |  |  |
| Tumor location | 1.04 (0.686–1.575) | 0.854 |  |  |
| (upper) |  |  |  |  |
| Basic disease | 1.132 (0.824–1.553) | 0.445 |  |  |

|  |  |  |
| --- | --- | --- |
| HBP | 1.02 (0.72–1.447) | 0.910 |
| DM | 1.325 (0.777–2.259) | 0.301 |
| CHD | 1.201 (0.589–2.449) | 0.614 |
| COPD | 0.541 (0.222–1.319) | 0.177 |

Abbreviations: BMI= Body Mass Index; Low-BMI < 18.5 kg/m<sup>2</sup>, 18.5 kg/m<sup>2</sup> ≤ Normal-BMI < 25.0 kg/m<sup>2</sup>, 25.0 kg/m<sup>2</sup> ≤ High-BMI; HBP= high blood pressure; DM= diabetes mellitus; COPD= chronic obstructive pulmonary disease; CHD= coronary heart disease; HR= hazard ratio; CI= confidence interval

There was a significant difference in OS between patients with cStage II and cStage III. Among the 46 patients with cStage II, the 3-year survival rate of the S group was 72%, while that of the NS group was 42% (HR, 0.351; 95%CI, 0.122–1.005; P=0.014; Figure 3A). Among the cStage III patients, the 1-year, 2-year, and 3-year OS rates of 84 patients in the S group were 79%, 62%, and 56%, respectively. The 1-year, 2-year, and 3-year OS rates of 56 patients in the NS group were 58%, 39%, and 36%, respectively (HR, 0.557; 95%CI, 0.333– 0.886; P=0.009; Figure 3B). No significant difference between the two groups was observed in cStage IV (Figure 3C).

**Figure 3.** Overall survival curves according to staging. (A) The overall survival curve between the S group and NS group(stage II); (B) The overall survival curve between the S group and NS group(stage III); (C) The overall survival curve between the S group and NS group (stage IV); (D) The overall survival curve between the S group and NS group after PSM (stage II); (E) The overall survival curve between the S group and NS group after PSM in stage III; (F) The overall survival curve between the S group and NS group after PSM (stage IV). PSM, propensity score matching.

In addition, patients with or without comorbidities were analyzed in the subgroups. For the patients with comorbidities, the 3-year survival rate was 58% for the S group and 19% for the NS group (HR, 0.312; 95%CI, 0.192–0.508; P<0.001; Figure 4A). In patients without comorbidities, the OS of the S group was significantly better than that of the NS group (HR, 0.521; 95%CI, 0.337–0.807; P=0.002; Figure 4B).

**Figure 4.** Overall survival curves for patients with and without comorbidities. (A) The overall survival curve between the S group and NS group for patients with a comorbidity; (B) The overall survival curve between the S group and NS group for patients without a comorbidity; (C) The overall survival curve between the S group and NS group for patients with a comorbidity after PSM; (D) The overall survival curve between the S group and NS group for patients without a comorbidity after PSM. PSM, propensity score matching.

To examine whether patients undergoing esophagectomy need a sufficient number of lymph node (LN) dissections, patients were divided into two subgroups: those who had ≥20 and <20 LNs dissected; the median number of resected LNs in the surgery group was 19. The OS was highest in the subgroup with ≥20 LN dissections, followed by the subgroups with <20 LN dissections and the NS group. The OS of patients in the subgroup with ≥20 LNs dissected was significantly better than that in the NS group (P<0.001). The OS of patients in the subgroup with <20 LNs dissected was still better than that in the NS group (P=0.020; Supplementary Figure 2A), The OS curve between the subgroups LNs ≥20 and LNs <20 dissected after PSM was calculated (P=0.060; Figure 5).

**Figure 5.** Overall survival curves. (A) The overall survival curve between the S subgroups and NS group. (B) The overall survival curve between the S subgroups and NS group after propensity score matching.

### 3.4. Adverse events of surgery, chemotherapy, and radiotherapy

The majority of adverse events were recorded one week after surgery, while anastomotic stenosis and delayed anastomotic fistula were generally recorded approximately one month after surgery. Adverse events in the S group mainly included pleural effusion in 55 patients (37.2%) and pulmonary infection in 31 patients (21.1%). In the NS group, gastrointestinal reactions (anorexia in 100 patients [73.5%], esophagitis in 51 patients [37.5%], and emesis in 37 patients [27.2%]) and hematologic toxicity (anemia [93 cases], leukocytopenia [84 cases], neutropenia [38 cases], and thrombocytopenia [37 cases]) were the most common adverse events. The number of adverse events in the NS group was higher than that in the S group (P<0.001), and they occurred mainly in patients with grades I–II (P<0.001). There was no significant difference in the number of patients with grades III–IV (P=0.679) and in those who died within 90 days of treatment (P=0.668) (Table 3). In terms of adverse events between the ≥20 and <20 LNs dissected subgroups of the S group (Table 3), which was based on the Clavien–Dindo classification (15-17), there were no significant differences between the two subgroups, whether in grades I–II (P=0.853), grade III (P=0.297), grade IV (P=0.470), or in total (P=0.139). In the S and NS groups, the mortality rate at 30 days was 1.4% and 1.5%, at 90 days was 4.7% and 5.9%, and at 6 months was 10.14% and 26.47%, respectively (Supplementary Table 1).

**Table 3.**
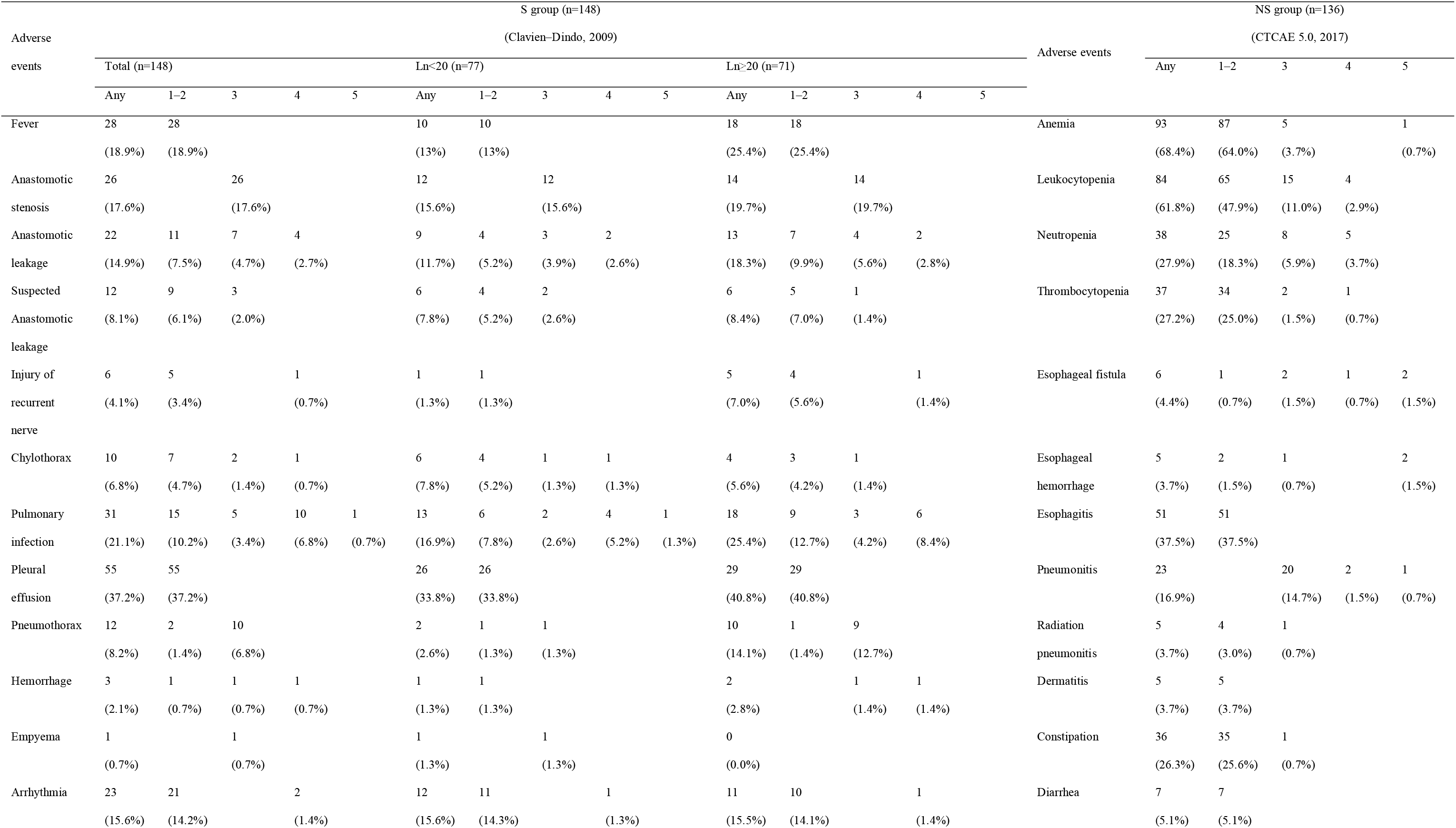
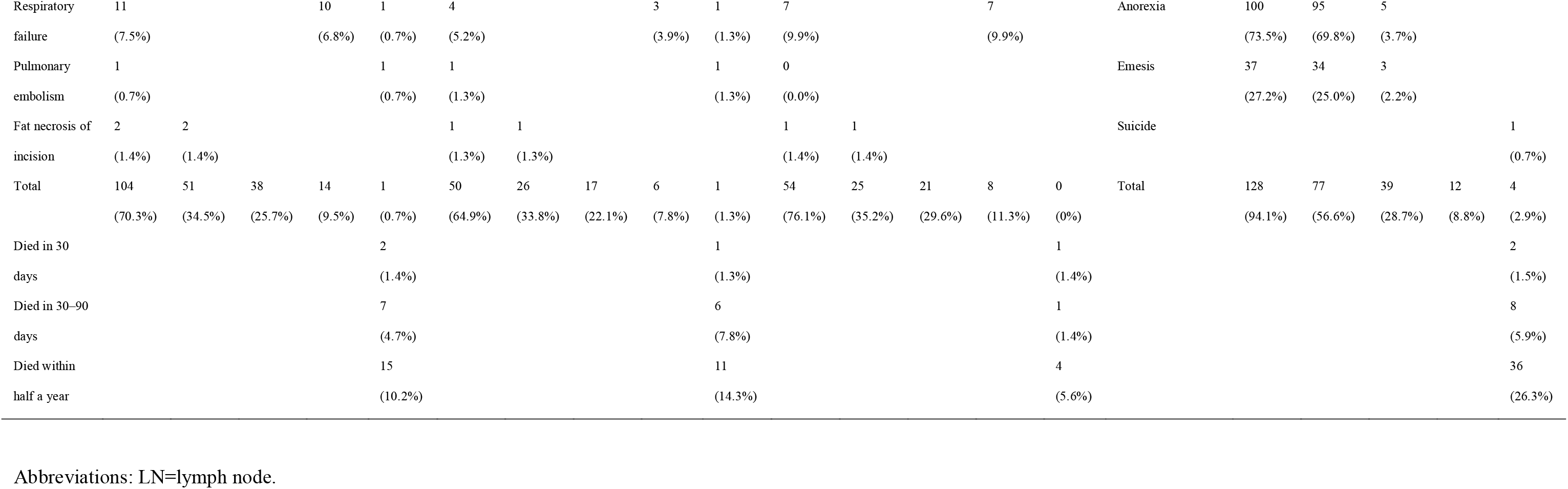
Adverse events in the surgical 8 and non-surgery groups.

### 3.5. Outcomes after PSM

After PSM, we found no significant difference in the basic characteristics of patients between the S group and NS group (Table 1). The median OS of the 140 patients was 23.8 months (95%CI, 11.3–36.3). The OS of the S group did not reach the median OS, and the average OS was 29.8 months (95%CI, 26.1–33.7). In contrast, the median OS in the NS group was only 16.8 months (95%CI, 8.8–24.7). The 1-year, 2-year, and 3-year OS rates in the S group were 76%, 61%, and 56%, respectively. In the NS group, the 1-year, 2-year, and 3-year OS rates were 56%, 38%, and 34%, respectively. The 1-year, 2-year, and 3-year OS rates were significantly lower in the NS group (HR, 0.553; 95%CI, 0.354–0.863; P=0.008; Figure 2C). Similarly, after excluding patients who died within 90 days of treatment, there were still significant differences in OS between the two groups (HR, 0.517; 95%CI, 0.323– 0.827; P=0.005; Supplementary Figure 4B). The 1-year, 2-year, and 3-year PFS rates in the S group and NS group were 73%, 54%, and 50%, and 52%, 34%, and 29%, respectively (HR, 0.629; 95%CI, 0.417–0.951; P=0.024; Figure 2D).

As for clinical staging of the 24 cStage II patients, the OS of the N group was better than that of the NS group. The 3-year OS rates in the S and NS groups were 83% and 36% respectively (HR, 0.206; 95%CI, 0.063–0.671; P=0.014; Figure 3D). There were 94 patients in cStage III; 46 in the S group and 40 in the NS group. The 1-year, 2-year, and 3-year OS rates in the S group were 83%, 63%, and 57%, respectively. In the NS group, the 1-year, 2-year, and 3-year OS rates were 58%, 39%, and 36%, respectively (HR, 0.583; 95%CI, 0.298– 0.971; P=0.033; Figure 3E). No significant difference in OS was observed between the N and NS groups among patients with cStage IV (HR, 0.793; 95%CI, 0.377–1.861; P=0.583; Figure 3F).

In patients with comorbidities, the 3-year OS rates of the S and NS groups were 60% and 25%, respectively (HR, 0.406; 95%CI, 0.206–0.800; P=0.008; Figure 4C). However, the difference between the two groups of patients without comorbidities was not obvious after PSM (HR, 0.682; 95%CI, 0.378–1.233; P=0.196; Figure 4D). Survival differences were only observed between the subgroup with LNs ≥20 and the NS group (P=0.004; Figure 5).

Adverse events in the S group mainly included pleural effusion in 30 (42.6%) patients and pulmonary infection in 14 (20.0%) patients. Mainly grade 1 to 3 adverse events occurred, of which grade 1 to 2 adverse events accounted for 41.4%, of the events and grade 3 adverse events accounted for 22.9%. In the NS group, 65 patients had adverse reactions. Anemia occurred in 47 (67.1%), leukopenia in 41 (58.6%), and anorexia in 48 cases (68.6%). Grade 1 to 2 adverse events accounted for 57.1% of the events and grade 3 adverse events accounted for 25.7%. The number of adverse events in the NS group was higher than that in the S group (P=0.002) (Table 3).

## 4. DISCUSSION

In this study, we discussed various treatments for older adult patients aged 70 years or older with ESCC and compared esophagectomy and conservative treatment in terms of OS, PFS, and complications after treatment. Patients with ESCC older than 70 years who underwent esophagectomy had significantly better OS and PFS than those who underwent non-surgical treatment based on RT and/or CT.

In a Japan Clinical Oncology Group phase III trial (JCOG 9907) to determine the optimal timing for CT, the overall frequency of adverse events was 34%. Recurrent nerve palsy (18%) and pneumonia (13%) were the main adverse events. In the JCOG 1109 NExT study, the frequency of pneumonia was 10.3%, 9.8%, and 12.9% and the frequency of recurrent nerve palsy was 15.1%, 8.7%, and 12.4% in the Neo CF, Neo DCF, and Neo CF+RT groups, respectively [26,27]. However, in our study, the frequency of adverse events was 70.3% in the S group and 94.1% in the NS group, the frequency of pulmonary infection was 21.1% in the S group, and the frequency of pneumonitis was 16.9% in the NS group. Recurrent nerve injury occurred in 4.1% of cases, which was similar to results from the NEOCRTEC5010 study [2]. Furthermore, adverse events did not significantly differ between the S group and the NS group, when the patients had grade III–IV ESCC. However, patients experienced less adverse events when they exhibited grades I–II ESCC. The results after PSM were similar to those before PSM. To explore the reasons for the different OS outcomes after PSM, we conducted a subgroup analysis. Patients with TNM stages II, III, or IV had the same results after PSM; in TNM stage II or III, the patients’ OS was better in the surgical treatment group. Nevertheless, OS did not differ significantly between the S and NS groups when patients had ESCC stage IV. The results indicate that accurate staging and appropriate adjustment of the treatment plan for different patients are critical.

The study findings suggest that the ideal treatment strategy for patients with ESCC aged 70 years or older should be esophagectomy. However, older adults are prone to surgical complications, and adverse events due to RT or CT are also prominent. Older adult patients have more complications in the peri-chemoradiotherapy period, a lower long-term OS rate, and poor tolerance to this treatment modality, which was also confirmed in the 2010 study at the Massachusetts General Hospital and the 2020 study at the Acharya Tulsi Regional Cancer Treatment and Research Institute [28,29]. Despite adverse events during therapy, radical treatment remains a necessity according to a 2017 study by the Affiliated Hospital of Guizhou Medical University. In patients aged >75 years with esophageal cancer, CT combined with RT was more advantageous than RT alone [30]. In a study from the Changhua Christian Hospital, in patients with early esophageal squamous cell carcinoma, esophagectomy alone had a significant advantage in terms of survival over definitive CRT. However, among patients with locally advanced disease, the overall survival was similar between the two treatment modalities [31]. However, lymphadenectomy might be considered as an important factor vital to OS, according to recent studies and the present data [32–33]. A Turkish study compared the OS of patients who underwent surgery as opposed to CRT and showed no significant differences in patients with locally advanced esophageal cancer [34]. However, a Japanese study which used PSM showed the opposite outcome [35]. Our study showed that surgical treatment brought significant improvements in the OS of patients.

In the well-known phase III study, FFCD 9102, the 2-year survival rate of patients treated with neoadjuvant CRT combined with surgery and CRT alone was approximately 30% [36]. As models of esophageal cancer treatments continue to evolve, differences in treatment patterns are emerging. Therefore, we collected data from prospective databases from January 2018 to September 2019 and found through subgroup analysis that among patients with comorbidities, there were no significant differences between the S and NS groups after PSM. Therefore, the comorbidities of patients should also be considered before surgical treatment. Furthermore, patients with no less than 20 resected LNs had better OS than those with less than 20 resected and the same results were obtained after PSM.

In the current treatment models of ESCC, comprehensive treatment based on surgery is still predominant [2−5,26,27]. Neoadjuvant treatment modalities also are advantageous for patients, as shown by a meta-analysis from Germany which compared neoadjuvant treatment modalities and definitive non-surgical therapy based on radiation or CT for ESCC. Neoadjuvant treatment modalities significantly improved OS without increasing morbidity in patients with resectable ESCC. Definitive non-surgical therapy did not show any survival benefit compared to the neoadjuvant mode [37]. Regarding the optimal treatment for older adult patients with ESCC, further international multicenter studies with a large sample size are needed to establish the optimal therapy in future.

This study has some limitations. First, this is a retrospective study; hence, the required data were often missing and difficult to verify, while the results were biased. Second, as there were many different models of conservative treatment used, therapeutic effects on the OS of patients were heterogeneous; thus, bias was unavoidable in the results. Third, data were collected from a single-center database, and lack of 5-year OS and PFS. With the development of immunotherapy in ESCC, multicenter prospective phase III clinical trials employing the advanced non-surgical treatment options are required to further clarify the treatment of patients aged >70 years.

## 5. CONCLUSIONS

Patients with ESCC older than 70 years who underwent esophagectomy had significantly better OS and PSF than those who underwent non-surgical treatment based on RT and/or CT. Regarding adverse events, patients who underwent esophagectomy had advantages in the reduced number of adverse events compared to patients who underwent non-surgical treatments based on RT and/or CT.

## Data Availability

All data produced in this retrospective study was reviewed by the Ethics Committee (EC) for Medical Research and New Medical Technology of Sichuan Cancer Hospital (SCCHEC-02-2022-050). The data are anonymous, and the requirement for informed consent was therefore waived.

## ACKNOWLEDGEMENTS

We appreciate Yongtao Han, Xuefeng Leng, Qifeng Wang, and Chenghao Wang for their financial support and everyone who contributed to the data collection.

## DISCLOSURES

### Ethics information

All procedures performed in this study were in accordance with the Declaration of Helsinki (as revised in 2013). This study was approved by the Ethics Committee of the Sichuan Cancer Hospital (No. NCC2014ZC-01) and Sichuan Science and Technology Project (No. 2021YJ0118) and this retrospective study was reviewed by the Ethics Committee (EC) for Medical Research and New Medical Technology of Sichuan Cancer Hospital (SCCHEC-02-2022-050). The data are anonymous, and the requirement for informed consent was therefore waived.

### Previous communication

Partial preliminary results of this article were poster presented by the first author of this article at 31th European Conference on General Thoracic Surgery (ESTS 2023) which will held virtually from 4-6 June 2023.

## Author contributions

Study concept and design: All authors. Acquisition, analysis, or interpretation of data: All authors. Drafting of the article: Kexun Li. Revising the article critically for important intellectual content: All authors. Final approval of the version to be published: All authors. Statistical analysis: Changding Li. Obtained funding: Lin Peng. Administrative, technical, or material support: Yongtao Han, Xuefeng Leng and Lin Peng. Study supervision: Xuefeng Leng and Qifeng Wang. Kexun Li, Changding Li, and Xin Nie had full access to all the data in the study and take responsibility for the integrity of the data and the accuracy of the data analysis. The work reported in the paper has been performed by the authors, unless clearly specified in the text.

## Provenance and peer review

Not commissioned, externally peer-reviewed.

## REFERENCES

1 Sung H, Ferlay J, Siegel RL, et al. Global Cancer Statistics 2020: GLOBOCAN Estimates of Incidence and Mortality Worldwide for 36 Cancers in 185 Countries. CA A Cancer J Clin 2021;71:209–49. doi:10.3322/caac.21660

2 Yang H, Liu H, Chen Y, et al. Neoadjuvant Chemoradiotherapy Followed by Surgery Versus Surgery Alone for Locally Advanced Squamous Cell Carcinoma of the Esophagus (NEOCRTEC5010): A Phase III Multicenter, Randomized, Open-Label Clinical Trial. JCO 2018;36:2796–803. doi:10.1200/JCO.2018.79.1483

3 Kelly RJ, Ajani JA, Kuzdzal J, et al. Adjuvant Nivolumab in Resected Esophageal or Gastroesophageal Junction Cancer. N Engl J Med 2021;384:1191–203. doi:10.1056/NEJMoa2032125

4 Shapiro J, Van Lanschot JJB, Hulshof MCCM, et al. Neoadjuvant chemoradiotherapy plus surgery versus surgery alone for oesophageal or junctional cancer (CROSS): long-term results of a randomised controlled trial. The Lancet Oncology 2015;16:1090–8. doi:10.1016/S1470-2045(15)00040-6

5 Li C, Zhao S, Zheng Y, et al. Preoperative pembrolizumab combined with chemoradiotherapy for oesophageal squamous cell carcinoma (PALACE-1). European Journal of Cancer 2021;144:232–41. doi:10.1016/j.ejca.2020.11.039

6 Yuan P, Xiang J, Borg M, et al. Analysis of lifetime death probability for major causes of death among residents in China. BMC Public Health 2020;20:1090. doi:10.1186/s12889-020-09201-7

7 National Cancer Institute: SEER cancer stat facts: esophageal cancer [online]. 2021. https://seer.cancer.gov/statfacts/html/esoph.html. (accessed 20 January 2021).

8 Vlacich G, Samson PP, Perkins SM, et al. Treatment utilization and outcomes in elderly patients with locally advanced esophageal carcinoma: a review of the National Cancer Database. Cancer Med 2017;6:2886–96. doi:10.1002/cam4.1250

9 Wright CD. Esophageal cancer surgery in 2005. Minerva Chir 2005;60:431–44.

10 Rentz J, Bull D, Harpole D, et al. Transthoracic versus transhiatal esophagectomy: A prospective study of 945 patients. The Journal of Thoracic and Cardiovascular Surgery 2003;125:1114–20. doi:10.1067/mtc.2003.315

11 Ozawa S, Ito E, Kazuno A, et al. Thoracoscopic esophagectomy while in a prone position for esophageal cancer: a preceding anterior approach method. Surg Endosc 2013;27:40–7. doi:10.1007/s00464-012-2404-3

12 Li K, Du K, Liu K, et al. Impact of two[field or three[field lymphadenectomy on overall survival in middle and lower thoracic esophageal squamous cell carcinoma: A single[center retrospective analysis. Oncol Lett 2023;25:189. doi:10.3892/ol.2023.13774

13 Motoori M, Ito Y, Miyashiro I, et al. Impact of Age on Long-Term Survival in Patients with Esophageal Cancer Who Underwent Transthoracic Esophagectomy. Oncology 2019;97:149–54. doi:10.1159/000500604

14 Motoyama S, Maeda E, Iijima K, et al. Does Esophagectomy Provide a Survival Advantage to Patients Aged 80 Years or Older? Analyzing 5066 Patients in the National Database of Hospital-based Cancer Registries in Japan. Annals of Surgery 2022;276:e16–23. doi:10.1097/SLA.0000000000004437

15 Van Deudekom FJ, Klop HG, Hartgrink HH, et al. Functional and cognitive impairment, social functioning, frailty and adverse health outcomes in older patients with esophageal cancer, a systematic review. Journal of Geriatric Oncology 2018;9:560–8. doi:10.1016/j.jgo.2018.03.019

16 Lagergren J, Bottai M, Santoni G. Patient Age and Survival After Surgery for Esophageal Cancer. Ann Surg Oncol 2021;28:159–66. doi:10.1245/s10434-020-08653-w

17 Da Costa BR, Cevallos M, Altman DG, et al. Uses and misuses of the STROBE statement: bibliographic study. BMJ Open 2011;1:e48. doi: 10.1136/bmjopen-2010-000048.

18 Stokes A, Collins JM, Grant BF, et al. Obesity Progression Between Young Adulthood and Midlife and Incident Diabetes: A Retrospective Cohort Study of U.S. Adults. Diabetes Care 2018;41:1025–31. doi:10.2337/dc17-2336

19 Reeves GK, Pirie K, Beral V, et al. Cancer incidence and mortality in relation to body mass index in the Million Women Study: cohort study. BMJ 2007;335:1134. doi:10.1136/bmj.39367.495995.AE

20 Low DE, Alderson D, Cecconello I, et al. International Consensus on Standardization of Data Collection for Complications Associated With Esophagectomy: Esophagectomy Complications Consensus Group (ECCG). Annals of Surgery 2015;262:286–94. doi:10.1097/SLA.0000000000001098

21 Remmelt Veen, Jan-Willem H. P. Lard M. Recording and Classification of Complications in a Surgical Practice. The European Journal of Surgery 1999;165:421–4. doi:10.1080/110241599750006622

22 Clavien PA, Sanabria JR, Strasberg SM. Proposed classification of complications of surgery with examples of utility in cholecystectomy. Surgery 1992;111:518–26.

23 Cunningham SC, Kavic SM. What Is a Surgical Complication? World J Surg 2009;33:1099–100. doi:10.1007/s00268-008-9881-5

24 Bruner DW, Hanisch LJ, Reeve BB, et al. Stakeholder perspectives on implementing the National Cancer Institute’s patient-reported outcomes version of the Common Terminology Criteria for Adverse Events (PRO-CTCAE). Behav Med Pract Policy Res 2011;1:110–22. doi:10.1007/s13142-011-0025-3

25 Ji Y, Du X, Zhu W, et al. Efficacy of Concurrent Chemoradiotherapy With S-1 vs Radiotherapy Alone for Older Patients With Esophageal Cancer: A Multicenter Randomized Phase 3 Clinical Trial. JAMA Oncol 2021;7:1459. doi:10.1001/jamaoncol.2021.2705

26 Kataoka K, Takeuchi H, Mizusawa J, et al. Prognostic Impact of Postoperative Morbidity After Esophagectomy for Esophageal Cancer: Exploratory Analysis of JCOG9907. Annals of Surgery 2017;265:1152–7. doi:10.1097/SLA.0000000000001828

27 Kato K, Ito Y, Daiko H, et al. A randomized controlled phase III trial comparing two chemotherapy regimen and chemoradiotherapy regimen as neoadjuvant treatment for locally advanced esophageal cancer, JCOG1109 NExT study. JCO 2022;40:238–238. doi:10.1200/JCO.2022.40.4_suppl.238

28 Mak RH, Mamon HJ, Ryan DP, et al. Toxicity and outcomes after chemoradiation for esophageal cancer in patients age 75 or older: ChemoRT, esophageal cancer in the elderly. Diseases of the Esophagus 2009;23:316–23. doi:10.1111/j.1442-2050.2009.01014.x

29 Mohata S, Kumar H, Sharma N, et al. Acute treatment-related toxicity in elderly patients with good performance status compared to young patients in locally advanced esophageal carcinoma treated by definitive chemoradiation: A retrospective comparative study. J Can Res Ther 2020;16:116. doi:10.4103/jcrt.JCRT_878_18

30 Zhao Q, Hu G, Xiao W, et al. Comparison of definitive chemoradiotherapy and radiotherapy alone in patients older than 75 years with locally advanced esophageal carcinoma: A retrospective cohort study. Medicine 2017;96:e7920. doi:10.1097/MD.0000000000007920

31 Wang B-Y, Hung W-H, Wu S-C, et al. Comparison Between Esophagectomy and Definitive Chemoradiotherapy in Patients With Esophageal Cancer. The Annals of Thoracic Surgery 2019;107:1060–7. doi:10.1016/j.athoracsur.2018.11.036

32 Li K, Leng X, He W, et al. Resected lymph nodes and survival of patients with esophageal squamous cell carcinoma: an observational study. International Journal of Surgery 2023;Publish Ahead of Print. doi:10.1097/JS9.0000000000000436

33 Li K, Nie X, Li C, et al. Mapping of Lymph Node Metastasis and Efficacy Index in Thoracic Esophageal Squamous Cell Carcinoma: A Large-Scale Retrospective Analysis. Ann Surg Oncol Published Online First: 25 May 2023. doi:10.1245/s10434-023-13655-5

34 Sakin A, Sahin S, Aldemir MN, et al. Chemoradiotherapy followed by surgery versus observation in esophageal squamous cell carcinoma. J BUON 2021;26:1509–16.

35 Nomura M, Oze I, Kodaira T, et al. Comparison between surgery and definitive chemoradiotherapy for patients with resectable esophageal squamous cell carcinoma: a propensity score analysis. Int J Clin Oncol 2016;21:890–8. doi:10.1007/s10147-016-0963-3

36 Bedenne L, Michel P, Bouché O, et al. Chemoradiation Followed by Surgery Compared With Chemoradiation Alone in Squamous Cancer of the Esophagus: FFCD 9102. JCO 2007;25:1160–8. doi:10.1200/JCO.2005.04.7118

37 Kranzfelder M, Schuster T, Geinitz H, et al. Meta-analysis of neoadjuvant treatment modalities and definitive non-surgical therapy for oesophageal squamous cell cancer. British Journal of Surgery 2011;98:768–83. doi:10.1002/bjs.7455

